# Exome Analysis Points *APOE4* Haplotype as Major Risk to Develop Mesoamerican Nephropathy

**DOI:** 10.1101/2024.02.22.24303190

**Authors:** Iván Landires, Karen Courville, Gumercindo Pimentel-Peralta, Raúl Cumbrera, Norman Bustamante, Mauricio Arcos-Burgos, Virginia Núñez-Samudio

**Affiliations:** Unidad de Genética y Salud Pública, Instituto de Ciencias Médicas, Las Tablas 0710-00043, Panama; Sistema Nacional de Investigación, Secretaría Nacional de Ciencia, Tecnología e Innovación, Ciudad del Saber, Clayton, Panama; Consulta de Genética Médica, Hospital Joaquín Pablo Franco Sayas, Ministry of Health, Las Tablas, Panama; Unidad de Hemodiálisis, Departamento de Nefrología, Hospital Dr. Gustavo N. Collado, Caja de Seguro Social, Chitré, Herrera, Panama; Grupo de Investigación en Psiquiatría (GIPSI), Departamento de Psiquiatría, Instituto de Investigaciones Médicas, Facultad de Medicina, Universidad de Antioquia, Medellín, Colombia

**Author notes:** These authors have contributed equally to this work and share senior authorship (Iván Landires, Karen Courville, Mauricio Arcos-Burgos, Virginia Núñez-Samudio). Correspondence to **Iván Landires, MD, PhD**, Instituto de Ciencias Médicas, Las Tablas, Los Santos, Panama. PO Box, 0710-00043.

## Abstract

The present study aims to characterize the genetic predisposition for the development of Mesoamerican Nephropathy (MeN) in patients from Panama. A Whole Exome Sequencing approach was performed in patients diagnosed with MeN using the criteria of the Pan American Health Organisation (PAHO), and clinical variables were analysed in all patients and in a group of exposed healthy donors and a group of non-exposed healthy donors. We found a variant located in the APOE gene (rs429358 c.388T>C p.(Cys130Arg), identified in 26% (16/61) of patients with MeN, that corresponds to the APOE4 haplotype. In the group of patients with the APOE4 haplotype we have identified that uric acid is elevated when compared to the group of patients without the haplotype. To the best of our knowledge, this is the first study to find variants in the APOE gene as an important genetic risk factor in patients with MeN. These patients have higher hyperuricemia than patients without the variant, which would indicate an important role of uric acid in the pathophysiology of the disease, suggesting the potential use of uric acid-lowering drugs in these patients.

## Introduction

The name Mesoamerican Nephropathy (MeN) defines a non-traditional Chronic Kidney Disease (CKDnt) with important morbi-mortality in Central America, observed in male and young workers exposed to metabolic and heat stress, whose primarily cause(s) remains elusive.^1^ The Pan American Health Organization (PAHO) defined patients with MeN as those with impaired renal function with a glomerular filtration rate (GFR) of less than 60 mL/min/m^2^, in the absence of predisposing factors of traditional CKD such as diabetes, hypertension, or glomerular disease.^2^ Chronic exposure to heavy metals, pesticides, working conditions with high temperatures and dehydration could increase the risk of developing MeN. A recent study identified that, in Panama, there is also an unusual increase in patients with MeN.^2^ MeN was found to be more prevalent among relatively young male patients in agricultural work activities exposed to pesticides. Hyperuricemia has also been identified as a significant clinical marker of MeN in patients from Panama.^2^

Genetic factors had remained mostly secluded from studies looking to dissect MeN’s etiology. Previous research using Genome Wide Association Studies (GWAS) in Sri Lanka and India have reported the association of genetic variants in the sodium-dependent dicarboxylate transporter member 3 (SLC13A3) and a voltage-gated K channel (KCN10A) with CKDnt.^3,4,5^ SLC13A3 gene encodes a transporter located in the basolateral membranes of human renal proximal tubules which could influence the development of the disease, although this remains to be investigated and clarified. KCNA10 is a potassium channel located in the glomerular endothelium and in the apical membrane of renal proximal tubular cells. Its presence in endothelial vascular smooth muscle cells may also regulate vascular tone, an intriguing mechanism that remains to be elucidated as to how it might influence the development of the disease. A recent study from Mexico using polymorphism genotyping has found variants in the gene (NOS3) that predispose to CKDnt through decreased renal nitric oxide production, leading to endothelial dysfunction and kidney damage in these patients.^6^

Exome sequencing has emerged as a first-line research and diagnostic method in kidney diseases.^7^

This study proposes for the first time the use of Whole Exome Sequencing (WES) to test the hypothesis of a probable genetic predisposition to MeN in panamanian patients.

## Methods

With a broad medical group of specialties: nephrologists, geneticists, epidemiologists, and bioinformaticians, we followed a cohort of 61 MeN patients, 30 exposed healthy donors to heat stress and agricultural work and 30 non-exposed healthy donors from the provinces of Herrera and Los Santos in Central Panama for more than five years diagnosed with a comprehensive clinical, laboratory, and genetics protocol (Whole Exome Sequencing and bioinformatic analysis, Human Ethics approval No. CIEI-CSS-M-181-12019 and funding from Panama’s “Secretaría Nacional de Ciencia, Tecnología e Innovación, SENACYT,” project IOMS19-013).

DNA extraction, preparation of libraries and next generation sequencing: Ten millilitres (10mL) of venous blood was drawn from the forearm vein of consenting study participants (EDTA tetra sodium anticoagulant). Samples were preserved at 4°C briefly before genomic DNA extraction using the QIAamp^®^ DNA Mini Kit (Qiagen, Hilden, Germany). DNA concentration was measured using Qubit 4™ Fluorometer (Thermo Fisher Scientific, USA). DNA samples with a concentration above 1.0ug were used to prepare sequencing libraries. Agilent liquid phase hybridization was applied to enrich whole exons. SureSelect Human All ExonV6 r2 (Agilent Technologies, CA, USA) was used for sequencing and capture libraries. Genomic DNA was randomly fragmented to 180-280bp using the Nextera DNA flex kit from Illumina, and then DNA fragments were end polished, A-tailed and ligated with the full-length adapter for Illumina sequencing. Fragments with specific indexes were hybridized with more than 543,872 biotin labelled probes after pooling, then magnetic beads with streptavidin were used to capture 334,401 exons from 20,967 genes. After PCR amplification and quality control, libraries were sequenced.

### Bioinformatics Analysis

All high-quality data were then mapped to the human genome assembly using the *bwa-mem* algorithm.^8^ The aligned files were processed with the Genome Analysis Tool Kit (GATK) to recalibrate base quality, realign indels and remove duplicates.^9^ This was followed by SNP and INDEL discovery and genotyping according to GATK Best Practices recommendations. All variant calls underwent quality score recalibration and filtering to remove low-quality variants. Genetic data were imported to Golden Helix^®^’s SVS 8.8.3 (Bozeman MT, USA), and quality control was performed, as recommended in several previous papers, ^10^ including specific criteria for CKD,^7^ using the following criteria: (*i*) fitting to Hardy-Weinberg equilibrium with *P*-values > 0.05/*m* (where *m* is the number of markers included for analysis); (*ii*) a minimum genotype call rate of 90%; (*iii*) and presence of two alleles, controlling for excess of heterozygosis by evaluating the departure from expected *f* (inbreeding-coefficient) values, *i*.*e*., significant deviation toward negative values of the *f coefficient*. Genotypic and allele frequencies were estimated by maximum likelihood. Variants with a minor allele frequency (MAF) ≥ 0.01 were classified as common and rare otherwise. Exonic variants with potential functional effect were identified using the annotations in the database for nonsynonymous SNPs’ functional predictions (dbNSFP, GRCh38/hg38 genome assembly). This filter uses PolyPhen-2, Provean, SIFT, Mutation Taster, Gerp^++^, PhyloP and Mutation Assessor to predict a variant’s deleterious effect and is fully implemented in the SVS 8.3.3 Variant Classification module. We also investigated the presence of variants associated to clinical disorders and annotated them according with the last report of ClinVar.

Clinical variables were compared between patients with and without genetic variants and between MeN patients and healthy donors.

## Results

We found a variant harbored in the APOE gene (rs429358 c.388T>C p.(Cys130Arg) belonging to the APOE4 haplotype, identified in 26% (16/61) of the patients, that exhibits an extraordinary and significant distortion of segregation (Hardy Weinberg Equilibrium of P= 0.001) when compared to unaffected populations from disparate regions around de world. At the first clinical assessment, initial blood uric acid levels in MeN patients (without any type of medical treatment) carrying the APOE4 haplotype had a significantly higher concentration than non-carriers individuals (Table 1). These results suggest that even within the group of patients with MeN, elevated serum uric acid is associated with an APOE-mediated genetic predisposition.

**Table 1.**
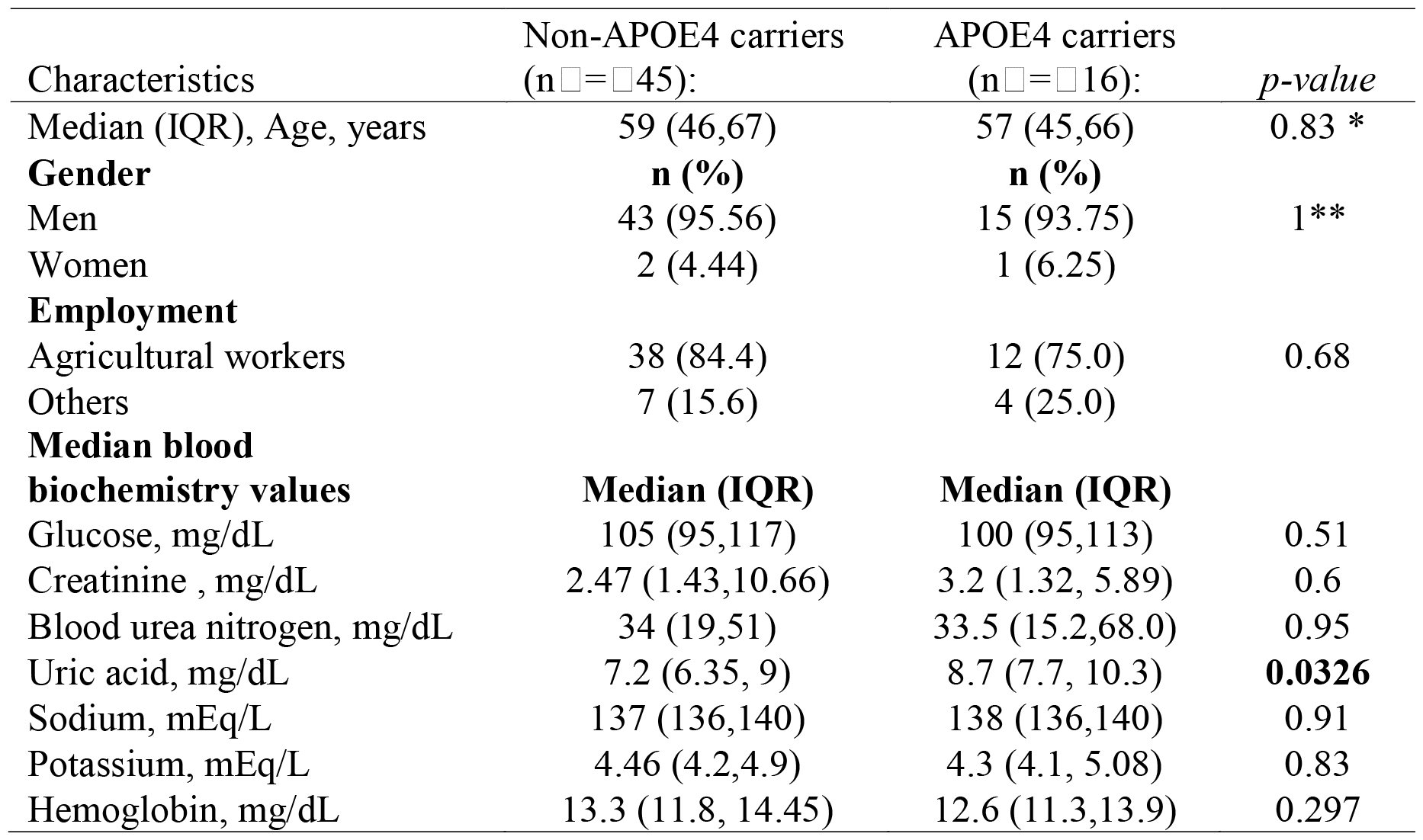
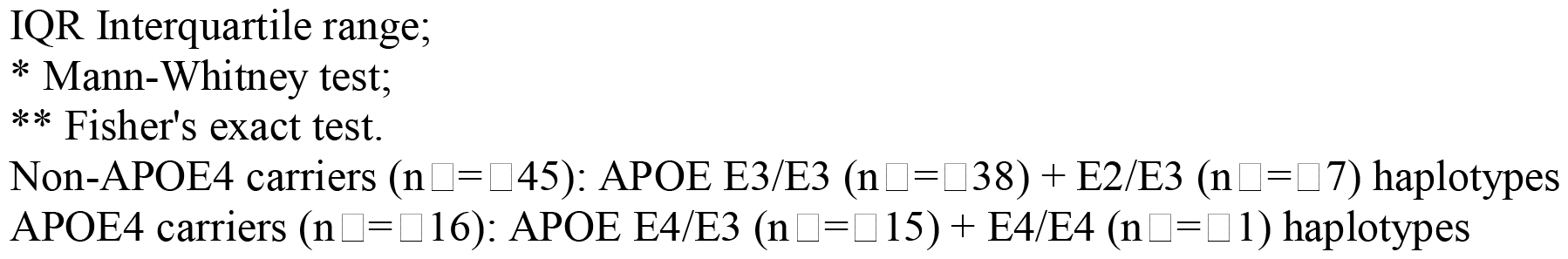
Clinical characteristics of MeN patients with APOE4 haplotype compared to non-carriers.

| Characteristics | Non-APOE4 carriers<br>(n=45): | APOE4 carriers<br>(n=16): | *p-value* |
| --- | --- | --- | --- |
| Median (IQR), Age, years | 59 (46,67) | 57 (45,66) | 0.83 * |
| **Gender** | **n (%)** | **n (%)** |  |
| Men | 43 (95.56) | 15 (93.75) | 1** |
| Women | 2 (4.44) | 1 (6.25) |  |
| **Employment** |  |  |  |
| Agricultural workers | 38 (84.4) | 12 (75.0) | 0.68 |
| Others | 7 (15.6) | 4 (25.0) |  |
| **Median blood**<br>**biochemistry values** | **Median (IQR)** | **Median (IQR)** |  |
| Glucose, mg/dL | 105 (95,117) | 100 (95,113) | 0.51 |
| Creatinine , mg/dL | 2.47 (1.43,10.66) | 3.2 (1.32, 5.89) | 0.6 |
| Blood urea nitrogen, mg/dL | 34 (19,51) | 33.5 (15.2,68.0) | 0.95 |
| Uric acid, mg/dL | 7.2 (6.35, 9) | 8.7 (7.7, 10.3) | **0.0326** |
| Sodium, mEq/L | 137 (136,140) | 138 (136,140) | 0.91 |
| Potassium, mEq/L | 4.46 (4.2,4.9) | 4.3 (4.1, 5.08) | 0.83 |
| Hemoglobin, mg/dL | 13.3 (11.8, 14.45) | 12.6 (11.3,13.9) | 0.297 |

IQR Interquartile range;
\* Mann-Whitney test;
\*\* Fisher's exact test.
Non-APOE4 carriers (n=45): APOE E3/E3 (n=38) + E2/E3 (n=7) haplotypes
APOE4 carriers (n=16): APOE E4/E3 (n=15) + E4/E4 (n=1) haplotypes

To follow these findings, we made a comparison with blood uric acid levels of a group of 60 MeN patients compared to those of 30 healthy controls exposed to high-temperature agricultural work and 30 unexposed healthy donors, finding significant differences between all these groups, highlighting that MeN patients showed the highest values, followed by the exposed healthy donors and then the unexposed healthy donors with the lowest values (Figure 1).

**Figure 1.**
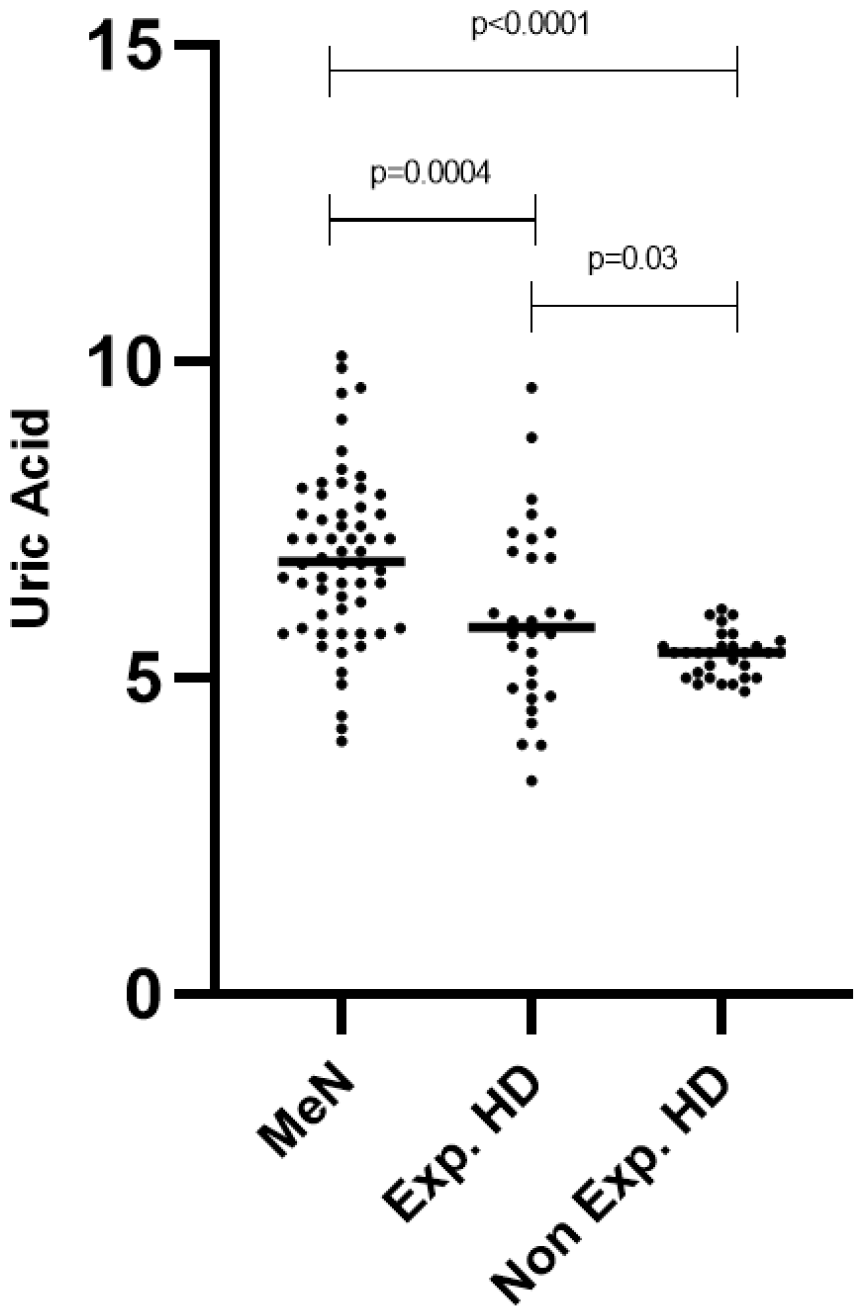
MeN: Mesoamerican Nephropathy; Exp.HD: Exposed healthy donors; Non Exp. HD: Non exposed healthy donors.

## Discussion

These results may be key findings, as previous studies by our group and others suggested that uric acid metabolic dysfunction after exercise and heat stress associated with dehydration-induced hyperuricemia is frequently observed in MeN patients.^2,11^ In earlier studies, the APOE4 has been associated with elevations in serum uric acid.^12^ Furthermore, previous studies have associated APOE variants with progression to CKD.^13^

Hyperuricemia is an independently predictive factor for CKD. Heat stress-associated urate crystalluria is frequent in subjects with MEN, in whom it may be favouring renal damage, and uricosuria accompanies rhabdomyolysis, in which it has been strongly suspected to play a contributing role. Uric acid has been shown experimentally to cause endothelial dysfunction by both depleting biologically active nitric oxide and inducing oxidative stress. Serum uric acid also is correlated with significantly elevated afferent arteriolar resistance in humans.^14^

We hypothesize that agricultural work at high temperatures elevates uric acid, even in healthy individuals, and that those with genetic predisposition, carriers of the APOE variant reported in this study, would be at increased risk of developing MeN. One limitation of the present study is the small sample of patients.

The link between APOE variation and the finding of hyperuricemia, both associated with MeN, points to the fact that uric acid dysfunction may be at the core of the pathophysiology of MeN, which is crucial for defining new treatments for hyperuricemia, such as uric acid-lowering therapies in healthy exposed individuals as well as in those with MeN.

## Data Availability

All data generated or analysed during this study are included in this published article.

## Acknowledgments

Iván Landires, Karen Courville and Virginia Núñez-Samudio are members of the Sistema Nacional de Investigación (SNI), which is supported by Panama’s Secretaría Nacional de Ciencia, Tecnología e Innovación (SENACYT).

## Authors’ contributions

Conceptualization, I.L., K.C., M.A.B., and V. N-S.; methodology, I.L., K.C., N.B., G.P.P., R.C., M.A.B., and V. N-S.; software, I.L., M.A.B., and V. N-S.; validation, I.L., K.C., M.A.B., and V. N-S.; formal analysis, I.L., K.C., G.P.P., R.C., M.A.B., and V. N-S.; investigation, I.L., K.C., N.B., G.P.P., R.C., M.A.B., and V. N-S.; resources, I.L., K.C., N.B., G.P.P., R.C., M.A.B., and V. N-S.; data curation, I.L., G.P.P., R.C., M.A.B., and V. N-S.; writing—original draft preparation, I.L., and V. N-S.; writing—review and editing, I.L., K.C., N.B., G.P.P., R.C., M.A.B., and V. N-S.; visualization, I.L., K.C., N.B., G.P.P., R.C., M.A.B., and V. N-S.; supervision, I.L., K.C., M.A.B., and V. N-S.;; project administration, I.L., and V. N-S.;; funding acquisition, I.L., K.C., and V. N-S.;; All authors have read and agreed to the published version of the manuscript.

## Availability of data and materials

All data generated or analysed during this study are included in this published article.

## Ethics approval and consent to participate

The study was conducted according to the guidelines of the

Declaration of Helsinki, and approved by the Interinstitutional Ethics Committee of the Social Security

Fund and the National Directorate for Teaching and Research (Human Ethics approval No. CIEI-CSS-M-181-12019). No further administrative permissions were needed to access the raw data used in this study. The data used in this study were anonymized before use.

